# The SARS-CoV-2 receptor angiotensin-converting enzyme 2 (ACE2) in Myalgic Encephalomyelitis/Chronic Fatigue Syndrome: analysis of high-throughput epigenetic and gene expression studies

**DOI:** 10.1101/2021.03.23.21254175

**Authors:** João Malato, Franziska Sotzny, Sandra Bauer, Helma Freitag, André Fonseca, Anna D Grabowska, Luís Graça, Clara Cordeiro, Luís Nacul, Eliana M Lacerda, Jesus Castro-Marrero, Carmen Scheibenbogen, Francisco Westermeier, Nuno Sepúlveda

## Abstract

Patients affected by Myalgic Encephalomyelitis/Chronic Fatigue Syndrome (ME/CFS) show specific epigenetic and gene expression signatures of the disease. However, it is unknown whether these signatures include abnormal levels of the human angiotensin-converting enzymes, ACE and ACE2, the latter being the main receptor described for the host-cell invasion by SARS-CoV-2. To investigate that, we first re-analyzed available case-control epigenome-wide association studies based on DNA methylation data, and case-control gene expression studies based on microarray data. From these published studies, we found an association between ME/CFS and 4 potentially hypomethylated probes located in the *ACE* locus. We also found another disease association with one hypomethylated probe located in the transcription start site of ACE2. The same disease associations were obtained for women but not for men after performing sex-specific analyses. In contrast, a meta-analysis of gene expression levels could not provide evidence for a differentially expression of *ACE* and *ACE2* in affected patients when compared to healthy controls. In line with this negative finding, the analysis of a new data set on the gene expression of *ACE* and *ACE2* in peripheral blood mononuclear cells did not find any differences between a female cohort of 37 patients and 34 age-matched healthy controls. Future studies should be conducted to extend this investigation to other potential receptors used by SARS-CoV-2. These studies will help researchers and clinicians to improve the understanding of the health risk imposed by this virus when infecting patients affected by this debilitating disease.

## 1. INTRODUCTION

On March 11^th^, 2020, the World Health Organization officially declared the world to be under the fast-spreading pandemic of the Coronavirus disease 2019 (COVID-19) caused by the severe acute respiratory syndrome coronavirus-2 (SARS-CoV-2). This pandemic came in the aftermath of two past outbreaks of severe acute respiratory infections caused by other two human beta coronaviruses: the 2002/2003 Severe Acute Respiratory Syndrome (SARS) pandemic caused by SARS-CoV-1 and the 2012 Middle East Respiratory Syndrome (MERS) caused by MERS-CoV [1]. Since then, research efforts have been made to identify the molecular receptors by which the diverse coronaviruses are able to invade human host cells. Until now, the strongest candidate receptor is the human angiotensin-converting enzyme 2 (ACE2) whose interaction with the viral spike glycoprotein (S1) serves as a viral entry into host cells [2–4]. This enzyme is highly expressed in different organs, including the lungs, heart, kidneys, and skin [5–8]. Molecularly, ACE2 counteracts the effect of the angiotensin-converting enzyme (ACE), which results in the control of the blood pressure and systemic vascular resistance [9]. Failure to balance the expression of these genes is expected to lead to hypertension and cardiovascular diseases. Perturbation in the ACE/ACE2 ratio has also been hypothesized as key to the development of COVID-19 [10]. In line with these expectations, patients with severe symptoms of COVID-19 tend to show baseline hypertension and other chronic heart conditions [11–13]. To explain these clinical observations, it was hypothesized that these individuals could be at a higher risk of developing COVID-19 due to an upregulated *ACE2* expression [14]. ACE2-deficient individuals also seem to be at a higher risk of COVID-19, because viral entry typically induces a downregulation of this enzyme, which ultimately affects its balance with ACE [15]. In addition, a recent meta-analysis of drugs that raise *ACE2* expression indirectly (i.e., ACE inhibitors) provided no statistical association between these drugs and the mortality rate of COVID-19 [16]. Given these disparate lines of evidence, it is important to identify well-defined clinical populations in which *ACE2* expression could be impaired. These clinical populations can then be used to investigate the role of this enzyme in SARS-CoV-2 infections. Patients with Myalgic Encephalomyelitis/Chronic Fatigue Syndrome (ME/CFS) represent a neglected clinical population due to a poor recognition and limited knowledge of the disease by health staff and the society [17,18]. ME/CFS is a chronic disease characterized by an unexplained but persisting fatigue and post-exertional malaise as the hallmark symptoms among other clinical manifestations [19,20], which has been even associated with long-lasting COVID-19 symptoms [21]. The etiology of the disease remains largely unknown, but many patients report an infection at their symptoms’ onset [22,23]. Patients often show high prevalence of cardiovascular and endothelial dysfunctions, such as orthostatic intolerance, impaired blood pressure variability and arrhythmia [24–29]. These patients also show features of an unbalanced immune system consistent with an autoimmunity origin of the disease [30]. This immune perturbation could be a possible explanation for the frequent viral infections or the high rate of flu-like symptoms reported by ME/CFS patients [31]. Interestingly, ACE levels were found to be elevated in about 80% of patients diagnosed with an old case definition of ME/CFS [32]. Such observation suggested this enzyme as a possible biomarker for the disease. As far as we know, this biomarker potential was not investigated in follow-up studies. In turn, little is known about the role of ACE2 in patients with ME/CFS.

In the last two decades, there was an explosion of high-throughput technologies that allowed to interrogate the association of a large numbers of genetic variations, epigenetic changes, and altered gene expressions with complex diseases. Such technological developments motivated the research community to investigate specific epigenetic and gene expression signatures in patients with ME/CFS [33–35]. These investigations generated large amounts of data in which the role of ACE and ACE2 could be specifically assessed. The present paper then aimed to re-analyze these existing data in terms of ACE/ACE2 axis. With this purpose, we focused on studies comparing patients with ME/CFS to healthy controls (case-control study design). To strengthen existing evidence, we also reported the gene expression of *ACE* and *ACE2* in a new cohort of women affected by ME/CFS and healthy controls.

## 2. MATERIALS AND METHODS

### 2.1 Angiotensin I converting enzymes 1 and 2 (ACE and ACE2)

Human ACE and ACE2 are two homologous enzymes sharing 41% protein identity and 61% sequence similarity [36]. In more detail, ACE is a protein comprising a total of 1,306 amino acids (isoform 1) encoded by the *ACE* gene located on the q23.3 region of the chromosome 17 (genomic coordinates: 63,477,061-63,498,380 or 61,562,184-61,599,209 in the reference genomes hg38 and hg19, respectively). ACE2 is instead a protein with 805 amino acids of length encoded by the *ACE2* gene located on the p22.2 region of the X chromosome (genomic coordinates: 15,561,033-15,602,148 or 15,579,156-15,620,271 in the reference genomes hg38 and hg19, respectively).

### 2.2 Diagnosis of ME/CFS

Since there is still no disease-specific biomarker, many case definitions of ME/CFS have been proposed over the years [37]. These case definitions are invariantly based on the symptoms reported by suspected patients and on the exclusion of known pathologies that could explain fatigue. To reduce heterogeneity between studies, we only considered data from studies using either the 1994 Centre for Diseases Control criteria [19] or the 2003 Canadian Consensus Criteria [20]. These criteria are hereafter denoted as 1994 CDC/Fukuda definition and 2003 CCC, respectively. Note that the choice of these two criteria is in line with the recent recommendations for research given by the European Network on ME/CFS [18].

### 2.3 Analysis of epigenome-wide association studies (EWAS)

We focused our analysis on four available case-control EWAS on ME/CFS [38–41], which were reviewed elsewhere [34], and two additional case-control studies published after this review [42,43] (Table 1). These studies were conducted using Illumina methylation arrays with the exception of a single study which used the reduced representation bisulfite sequencing technology [43] (Table 1). We conducted a joint analysis of the four array-based studies which had the data publicly available in the NCBI Gene Expression Omnibus (GEO) data repository [44]. For the remaining two studies, we analyzed the lists of significant differentially methylated probes between patients and healthy controls and checked whether these lists contained any probes located in the genes of interest. Our analysis of the array-derived data referred to the probes located in the coding regions and the transcription start sites (TSS) of *ACE* and *ACE2*, respectively. In addition, we restricted our analysis to the probes of the Infinium HumanMethylation450K array shared with the Infinium HumanMethylationEPIC array: 19 probes in *ACE* (chromosome 17) and 8 probes in *ACE2* (chromosome X; Supplementary Table 1). We performed an initial quality control of these probes based on (i) their probability of detection, (ii) their cross-reactivity with other genomic regions due to high sequence homology, and (iii) whether their location was associated with any single nucleotide polymorphism (SNP) [45]. Given that these four studies were conducted in European or North American populations, probes were considered problematic if the associated SNPs had a minor allele frequency higher than 0.05 in these populations (Supplementary Table 2). All probes were considered non-problematic and they were included in the analysis.

**Table 1.**
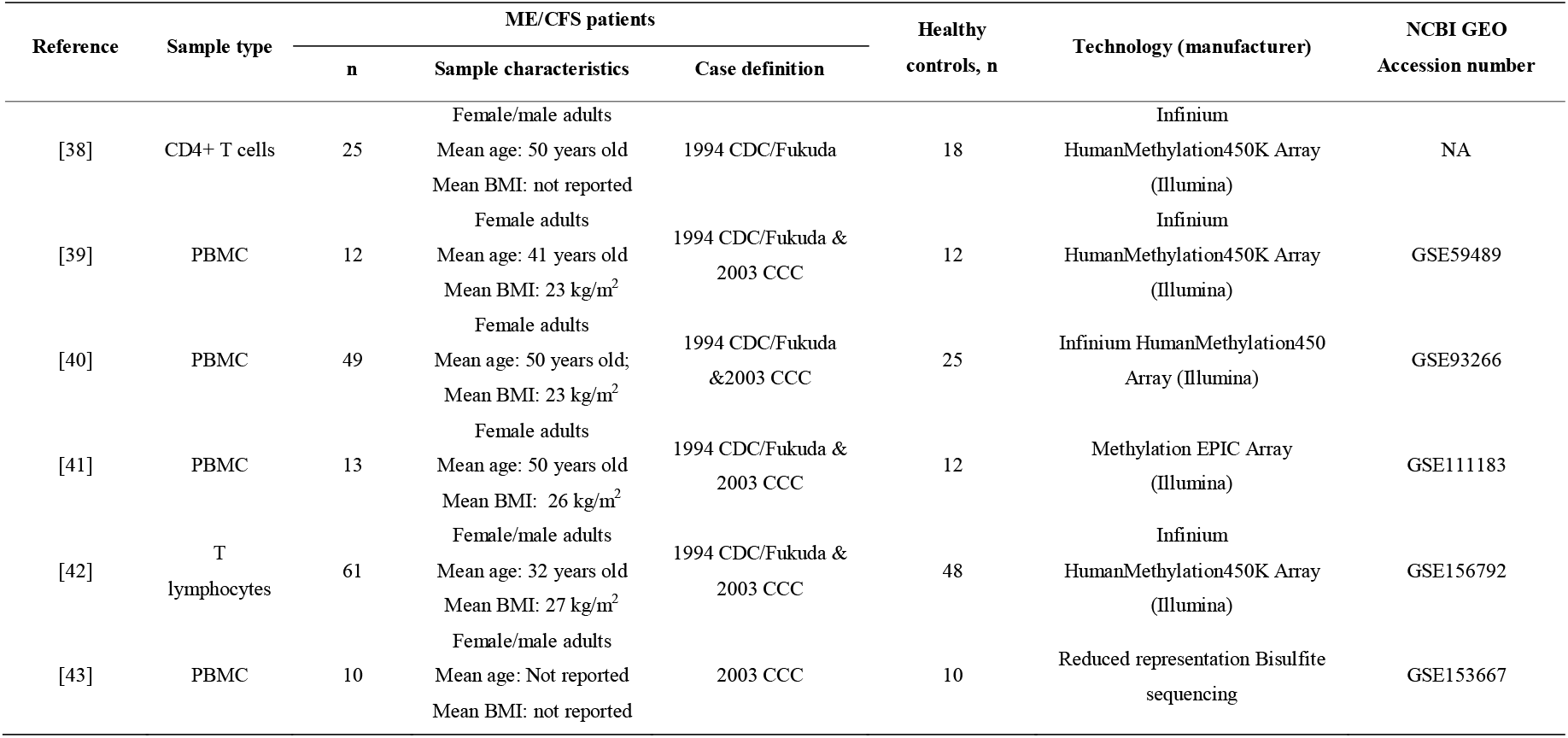
Summary of the six EWAS under analysis.

We then performed a joint analysis of the overall data from these array-based studies. Note that this analysis is equivalent to the so-called pooled individual-patient level data analysis that can be used for meta-analysis purposes [46]. For each data set, methylation signals were given by *β*-values (defined as the ratio between the methylated signal divided by the total of methylated and non-methylated signals). To obtain a good approximation of the data to the Normal distribution, *β-* values were converted into the corresponding M-values using the logit transformation [47]. To analyze data of each CpG probe, we initially fitted a linear regression model with the M-values as the outcome variable, and a study indicator variable and disease status as covariates. In this model, we considered the main effects plus the respective interaction terms. Note that, in this model, the main effect of the disease status can be seen as the pooled estimate across all studies as done in traditional meta-analyses. After performing parameter estimation, the model was then simplified using a backward stepwise procedure based on Akaike’s information criteria. Since the study indicator variable was always statistically significant when analyzing data from different probes, we reported the evidence for association of a given probe with ME/CFS by the p-value of a likelihood ratio test. In this test, we compared the model including the study indicator variable only with the best model including that covariate and disease status (i.e., either a model including main effects only or a model including the main effects plus the interaction term). To adjust for multiple testing, we applied the Benjamini-Hochberg procedure [48] with a false discovery rate of 5% under the assumption of independent tests. This assumption was assessed by estimating the Pearson’s correlation coefficient for data of all possible pairs of probes.

We also conducted a sex-specific analysis of the four array-based studies. Note that three of these studies recruited female participants only, whilst the fourth study included both men and women (Table 1). However, the data of this latter study available in the NBCI GEO did not include information about the gender of each study participant. To overcome this issue, we used the whole DNA methylation data to estimate the gender of each study participant using the function *getSex* of the R package *minfi* [49]. The estimated frequencies of men and women were in agreement with those reported in the original study. For the women-specific analysis, we performed the same association analysis as described above. For the men-specific analysis, we compared two regression models for the data of each probe: (i) one model including no covariates and (ii) another model including the disease status as the covariate. The comparison was done by a likelihood ratio test whose p-values were then adjusted for multiple testing as described above.

### 2.4 Analysis of gene expression studies (GES)

Our analysis was based on a total of eight case-control GES based on microarray technology, respectively (Table 2) [50–57]. These studies were based on PBMCs (5 studies), whole blood (2 studies) and muscle biopsies (one study). There were three additional case-control GES based on similar technology; however, these studies used unclear case definitions of ME/CFS [58,59] or case definitions other than 1994 CDC/Fukuda definition or 2003 CCC [60]. In addition, there were four case-control GES using RNA-seq technologies [61–64]. However, they were excluded from further analysis due to lacking of basic quality control checks, such as the percentage of reads that could be mapped onto the reference transcriptome, the percentage of the transcriptome covered, the average number of mapped reads per transcript, any putative effect of the GC content on the mapped read distribution, as recommended elsewhere [65].

**Table 2.** Summary of the 8 array-based GES under analysis, ordered by year of publication.

| Reference | Sample type | ME/CFS patients |  |  | Healthy controls, n | Technology (Manufacturer) | ACE/ACE2 available | Data availability (NCBI GEO Assession number) |
| --- | --- | --- | --- | --- | --- | --- | --- | --- |
|  |  | n | Sample characteristics | Case definition |  |  |  |  |
| [50] | PBMCs | 5 | Female adults<br>Mean age: 42 years old<br>Mean BMI: not reported | 1994<br>CDC/Fukuda | 5 | Atlas Glass Human 3.8 I<br>Microarray (BD<br>Biosciences Clontech) | No/No | No (NA) |
| [51] | PBMCs | 25 | Female/male adults<br>Mean age: 41 years old<br>Mean BMI: not reported | 1994<br>CDC/Fukuda | 25 | Custom microarray<br>(Nimblegen) | Unclear | No (NA) |
| [52] | Whole blood | 25 | Female/male adults<br>Mean age: 43 years old<br>Mean BMI: not reported | 1994<br>CDC/Fukuda | 50 | GeneChip Human<br>Genome U133 Plus 2.0<br>(Affymetrix) | Yes/ Yes | No (NA) |
| [53] | Whole blood | 11 | Female/male adults<br>Mean age: 34 years old<br>Mean BMI: 20.3 kg/m<sup>2</sup> | 1994<br>CDC/Fukuda | 11 | Custom microarray<br>(NA) | Yes/No | Yes (NA)<sup>a</sup> |
| [54] | Muscle biopsies | 4 | Female/male adults<br>Mean age: 45/37 years old<br>Mean BMI: not reported | 1994<br>CDC/Fukuda | 5 | Operon V2.0<br>(CRIBI University of<br>Padova) | Yes/Yes | No (NA) |
| [55] | PBMCs | 8 | Male adults<br>Median age: 36 years old<br>Mean BMI: not reported | 1994<br>CDC/Fukuda | 7 | GeneChip Human<br>Genome U133<br>(Affymetrix) | Yes/ Yes | Yes (GSE14577) |
| [56] | PBMCs | 37 | Female/male adults<br>Mean age: 51 years old<br>Mean BMI: 29.4 kg/m<sup>2</sup> | 1994<br>CDC/Fukuda | 25 | MWG 20K human Array<br>(Biotech MWG) | Yes/ Yes | No (NA) |
| [57] | PBMCs | 33 | Female/male adults<br>Mean age: not reported<br>Mean BMI: not reported | 1994<br>CDC/Fukuda | 21 | GeneChip Human Gene<br>ST (Affymetrix) | Yes/No | No (NA) |

<sup>a</sup> Data shared as a supplementary file in the online version of the study.

To analyze the selected microarray-based GES, we first searched for the annotation files associated with the technologies used. Most of these annotation files could be found in the NCBI GEO data repository [44]. We then used these annotation files to determine whether the respective microarrays included probes evaluating the expression of *ACE* and *ACE2*. We also checked whether each study made its data publicly available or at least reported any differential expression of *ACE* or *ACE2* genes between ME/CFS patients and healthy controls. As a qualitative assessment of the data, we compiled the information on whether each selected study performed any data normalization before inferring any differentially expressed genes.

A re-analysis was performed in studies in which any data normalization was conducted, had the respective expression data available, or at least reported differential expression of *ACE* or *ACE2* between patients and healthy controls. In studies where the available data did not resemble a Gaussian distribution in linear scale, we transformed the data by finding the optimal Box-Cox transformation. Since the resulting data resembled a Gaussian distribution, we calculated the classical t-based 95% confidence interval for the average difference between patients and healthy controls. This confidence interval was then converted back into a linear scale using the inverse of the optimal Box-Cox transformation used for the data. This confidence interval was finally log_2_-transformed in order to obtain the 95% confidence interval for the mean log_2_ fold change between patients and healthy controls. In studies in which the only available information was the mean log_2_ fold-change and the p-value associated with a Student’s t-test for testing differentially expressed genes, we determined the associated standard error and then calculated the 95% confidence interval for the mean log_2_ fold change. Finally, we pooled the different estimates of the mean log_2_ fold change using the inverse-weighted variance method for meta-analysis [66].

### 2.5 Analysis of new RNA data on the *ACE*/*ACE2* gene expression in ME/CFS

#### 2.5.1 Study participants

Thirty-seven female patients with ME/CFS were recruited in 2020 from the outpatient clinic for immunodeficiencies at the Institute for Medical Immunology at the Charité-Universitätsmedizin Berlin, Germany. Patients with ME/CFS were diagnosed according to the 2003 CCC while excluding other medical or neurological diseases which may cause fatigue [20]. Thirty-four female controls with self-reported healthy status were recruited from staff.

#### 2.5.2 Experimental procedure for RNA isolation and expression

PBMCs from study participants were isolated from heparinized whole blood by density gradient centrifugation using Biocoll Separating Solution (Merck Millipore). Total RNA was isolated from PBMCs (2×10^6^ cells) was extracted (NucleoSpin RNA Kit, Macherey-Nagel, cat. nr. 740955.50) according to the manufacturer’s instructions. Afterwards cDNA was prepared by reverse transcription (High-Capacity cDNA Reverse Transcription Kit, Applied Biosystems, cat. nr. 4368814) and real-time PCR was performed using TaqMan® Universal PCR Master Mix (cat. nr. 4305719) and TaqMan® Gene Expression Assays (cat. nr. 4331182) for *ACE* (Hs00174179_m1), *ACE2* (Hs01085333_m1) and the housekeeping gene *HPRT1* (Hs02800695_m1) (Applied Biosystems). For the amplification of *ACE* and *HPRT1* 20 ng and of *ACE2* 100 ng template cDNA were used. All measurements were performed with the ABI7200 and software Step One Plus as absolute quantification according to manufacturer’s instruction. Relative gene expression was analysed using the ΔCT method. Note that the expression of ACE2 mRNA was not possible to quantify for 11 patients due to insufficient cDNA material. Therefore, the analysis of *ACE2* gene expression was based on data from 26 patients and 34 healthy controls.

#### 2.5.3 Statistical analysis

We first tested whether the two groups were age-matched using the Kolgomorov-Smirnov test for two independent samples. For statistical convenience, raw gene expression data were independently transformed for ACE and ACE2 using the Box-Cox transformation. The estimates for the exponent of this transformation were 0.303 and 0.225 for ACE and ACE2, respectively. For each gene, a linear regression model was then applied to the resulting transformed data using age and disease status as covariates. The estimated linear regression models were then statistically validated by testing the normality assumption of the residuals using the Shapiro-Wilk test and by visually inspecting the assumption of constant variance of the same residuals as function of the covariates. The level of significance was set at 5% for this analysis.

After this analysis one could consider to pool estimates of fold-changes from this study with those obtained from previously published array-based GES. We did not attempt to perform such meta-analysis, because the nature of the data was very different between this study and these GES: RNA quantification by PCR versus intensity-based quantification, respectively.

#### 2.5.4 Ethical approval

The protocol of the German ME/CFS cohort study was approved by the Ethics Committee of Charité-Universitätsmedizin Berlin in accordance with the 1964 Declaration of Helsinki and its later amendments (reference number EA2/067/20). All patients and healthy controls recruited from staff gave written informed consent.

### 2.6 Statistical analysis and software

We performed our statistical analysis in the R software version 4.0.3. In this analysis, we used the following Bioconductor packages: *hgu133a*.*db, hgu133plus2*.*db, IlluminaHumanMethylation450kanno*.*ilmn12*.*hg19*, and *IlluminaHumanMethylationEPICanno*.*ilm10b2*.*hg19* to retrieve the annotation of the GeneChip HG-U133A, GeneChip U133+2, Infinium HumanMethylation450K Array and HumanMethylationEPIC arrays, respectively; *minfi* to estimate the sex of each individual from DNA methylation data [49].

## 3. RESULTS

### 3.1 Evidence from EWAS

In the six published EWAS, patients with ME/CFS were diagnosed using the 1994 CDC/Fukuda definition, the 2003 CCC, or both (Table 2). These patients were matched with healthy controls in terms of age, gender, and body mass index [39–41] with the exception of two studies where the matching was only based on the first two variables [38,43]. Trivedi et al [41] and Herrera et al [42] also matched for ethnicity, while the same matching could be assumed for the two other studies [39,40] given that these studies only recruited white females. Samples were derived from PBMCs [39–41], T lymphocytes [42], and CD4+ T cells [38]. Four of the six EWAS used the Infinium HumanMethylation450K Array by Illumina [38–40,42]. A single study was based on data generated from the Methylation EPIC Array [41]. Finally, the most recent study [43] used the reduced representation bisulfite sequencing.

The oldest EWAS [38] did not deposit the data in any public data repository and hence, our analysis of this study was only based on the reported 120 probes whose percentage of DNA methylation was significantly different between patients with ME/CFS and healthy controls. Although located in 70 known genes, these probes were neither located in *ACE* nor in *ACE2*.

The remaining four array-based EWAS [39–42] made the data publicly available and, therefore, we conducted a joint analysis of the respective data in accordance with a meta-analysis. With this purpose, we focused our analysis on the 27 probes available in both Infinium HumanMethylation450K and HumanMethylation EPIC arrays. These probes were not considered problematic in terms of co-hybridization with other genomic regions according to the list. Eight out of these 27 probes could be mapped onto genomic regions including SNPs within either *ACE* or *ACE2* genes (Supplementary Table 2). However, the associated SNPs were neither present in the European and North American populations nor had minor allele frequencies above 0.05 in the same populations (Figure 1A). Therefore, these probes were not considered problematic in this aspect.

**Figure 1.**
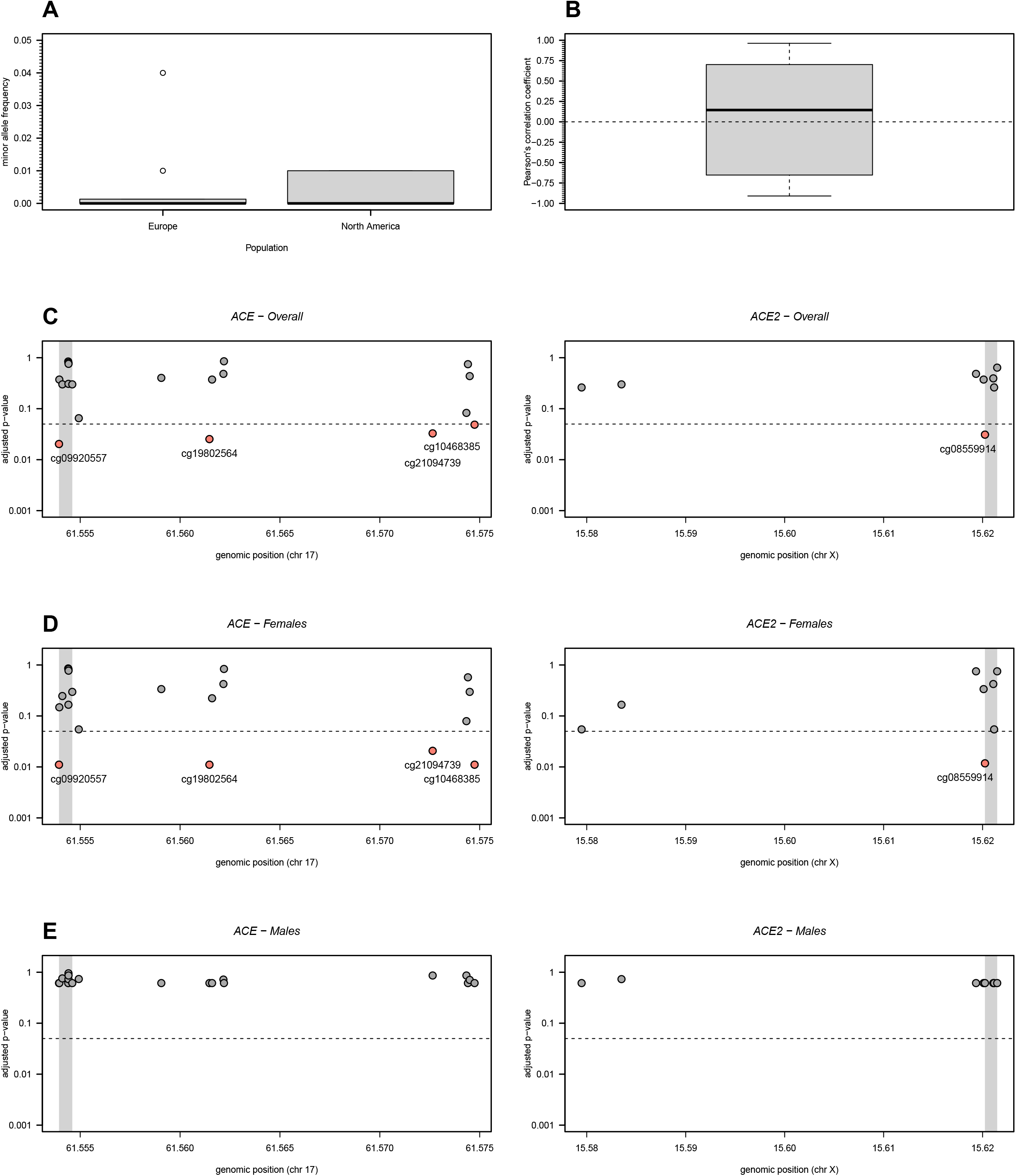
DNA methylation analysis of 19 and 8 CpG probes located in the *ACE* and *ACE2* genes, respectively. (**A**) Minor allele frequency in European and North American populations of SNPs located in the probes under analysis (see the respective data in Supplementary Table 2). (**B**) Boxplot of all possible Pearson’s correlation coefficients (y axis) between the M-values of the probes under analysis. Horizontal dashed line represents the situation of lack of correlation. (**C**) Adjusted p-values for the overall association between each probe and ME/CFS. Adjusted p-values were calculated according to the Benjamini-Hochberg procedure with a false discovery rate of 5% (dashed line). Grey areas in the plots represent the TSS of the genes. (**D**) and (**E**) The same analyses as shown in C but for women and men separately.

The joint analysis of these four studies revealed that the percentage of DNA methylation of the 27 probes of interest were on average uncorrelated with each other (Figure 1B). This observation supported the use of Benjamini-Hochberg procedure for controlling the overall false discovery rate given that this procedure assumes independent testing. The subsequent association analysis identified an association between ME/CFS and 4 CpG probes located in *ACE* (cg09920557, cg19802564, cg21094739, and cg10468385; Figure 1C). The probe cg09920557 is located in the TSS of the gene while the remaining probes are located in the gene body. The best linear regression models for these data contemplated not only the main effects of study and disease status but also the respective interaction (Supplementary Table 3). The interaction between study and disease could be visually seen when plotting the data (Figure 2A). Although not statistically significant, the estimated main effect of the disease status was negative for the analysis of each of these significant probes. This finding was in contrast with previous findings that patients with ME/CFS have a higher chance of possessing hypermethylated probes when compared to healthy controls [39,40].

**Figure 2.**
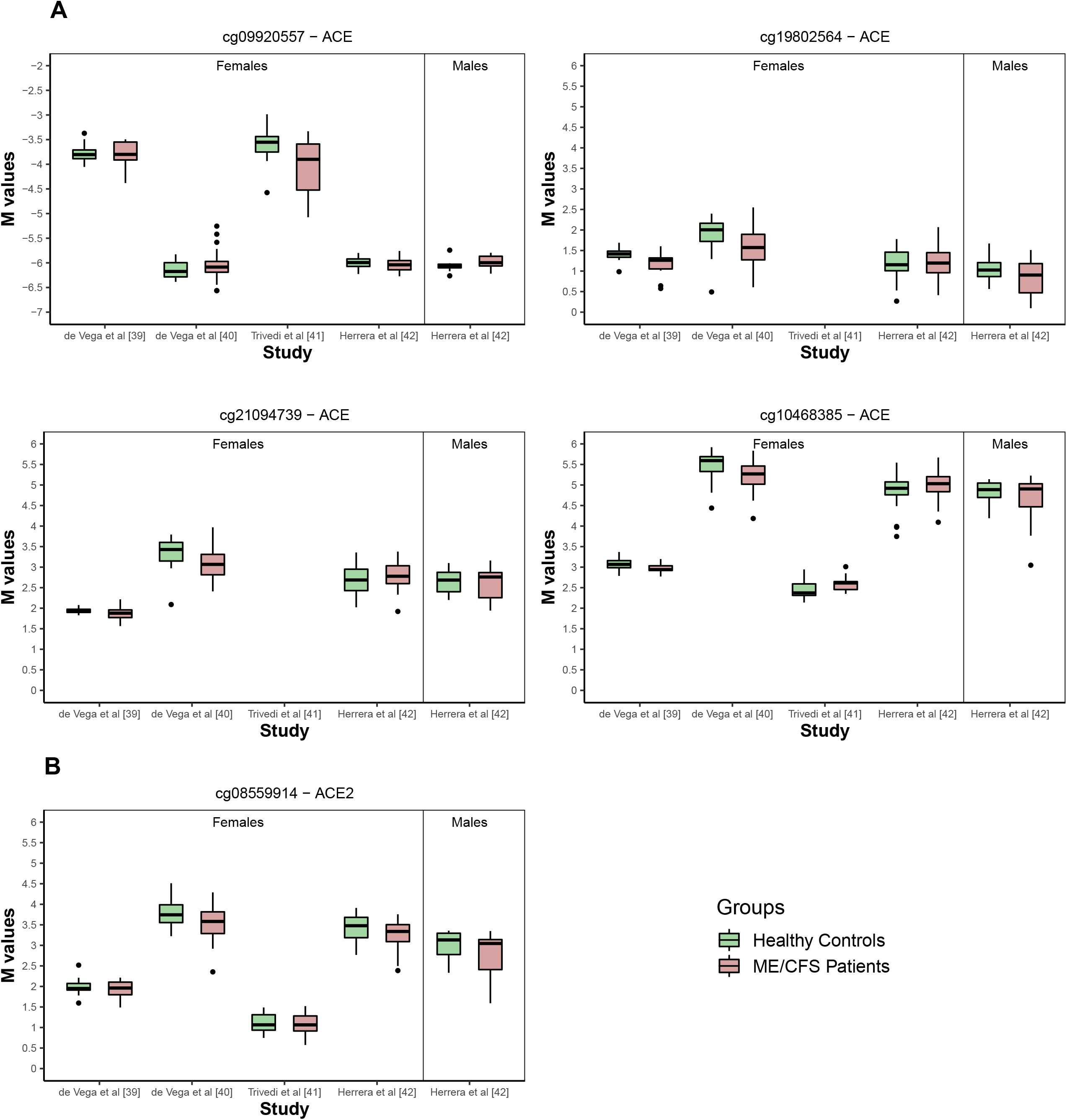
Boxplots per study, group and gender of the M-values referring to probes identified in Figures 1C and 1D. (**A)** Significant probes located in *ACE*. (**B**) Significant probe located in *ACE2*.

With respect to *ACE2*, the only significant association with ME/CFS was obtained for cg08559914, a probe located in the TSS of this gene (Figure 1C). According to the best model for the respective data, this probe is negatively associated with ME/CFS (coefficient estimate= −0.141 with a standard error of 0.048; Figure 2B and Supplementary Table 3). Given that the degree of methylation of the promoter regions is typically inversely correlated with gene expression, this finding suggested a putative increased expression of *ACE2* due to some hypomethylation of its TSS.

We then repeated the same association analysis but for women and men separately. For women, we obtained similar disease associations, as described for the overall analysis (Figure 1D and Supplementary Table 3). For men, we did not find any statistically significant associations probably due to limited data from a single study (Figure 1E). Therefore, the identified associations would appear to be specific to women.

Finally, the most recent EWAS was the only study that was not based on an array technology [43] and, as done above, our analysis consisted of analyzing the reported list of significant differentially methylated probes. This study reported 76 and 394 differentially methylated probes using two distinct statistical approaches for data analysis. These probes were located in 31 and 121 genes, respectively, which were neither *ACE* nor *ACE2* (see additional file 1 from Helliwell et al [43]).

### 3.2 Evidence from GES

The eight array-based GES under analysis were conducted in small cohorts of patients with ME/CFS (mean sample size=18.5; range=4-37) and healthy controls (mean sample size=18.6; range=5-50 individuals) (Table 3). In these studies, the patients and healthy controls were matched at least in terms of age and gender. Different commercial and custom microarray technologies were used for the respective gene expression quantification. There was only one study in which the microarray used did not include any probe in the genes of interest [50]. Another study used a custom array based on 9,522 genes from the RefSeq database as available in August 2002 [51].

**Table 3.** Summary statistics for the gene expression of *ACE* and *ACE2* from the German female study participants where data of *ACE2* were only available for 26 affected patients.

| Summary statistic | Healthy controls | ME/CFS patients |
| --- | --- | --- |
| N | 34 | 37 |
| Mean age (range), years | 37.4 (23; 65) | 41.1 (19; 60) |
| Mean disease duration<br>since diagnostic (range), months | NA | 5.4 (0; 24) |
| ACE |  |  |
| Geometric mean | 0.153 | 0.144 |
| Interquartile range | 0.087 | 0.073 |
| ACE2<sup>a</sup> |  |  |
| Geometric mean | 0.002 | 0.001 |
| Interquartile range | 0.005 | 0.004 |

However, it was unclear whether the gene expression of *ACE* and *ACE2* could have been quantified, because this study did not make available the list of genes included in the respective array. In terms of data sharing, only two studies made their data available either in the NCBI GEO data repository [55] or within the respective publication [53]. Although not sharing the data, there was a study that reported a significant association between ME/CFS and *ACE2* expression (log_2_(fold change)=0.190; 95% confidence interval=(0.021;0.359)) [56].

We conducted a re-analysis of the two studies in which the corresponding *ACE*/*ACE2* data was made available (Figure 3A). This analysis suggested a significant increase of *ACE* expression in patients with ME/CFS in data from Saiki et al [53] (mean log_2_(fold change)=0.470; 95% CI=(0.282;0.709)); this study used a custom array that consisted of stress-related genes not including *ACE2*. In opposition, the data from Gow et al [55] did not lead to any statistically significant result: −0.01 (95% CI=(−0.09;0.07)) and 0.00 (95% CI=(−0.08;0.07)) for probes 1 and 2 in the *ACE* gene, respectively; 0.04 (95% CI=(−0.12;0.19)) and 0.04 (95% CI=(−0.07;0.15)) for probes 1 and 2 in the *ACE2* gene, respectively (Figure 3A).

**Figure 3.**
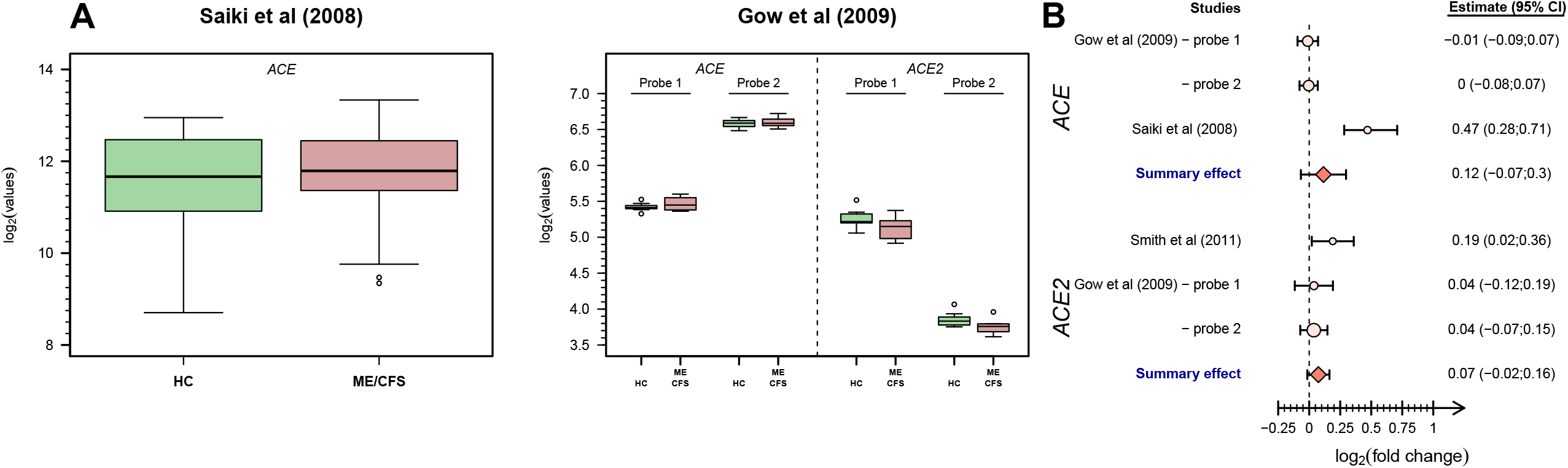
Analysis of *ACE*/*ACE2*-related data from eligible microarray-based GES. (**A)** Boxplots of the data from studies based on microarray technology. (**B**) Forest plot for the study-specific and pooled estimate of the mean log_2_ fold change between patients with ME/CFS and healthy controls using data shown in A.

To increase the overall statistical power to detect putative differentially expressed genes, we pooled the estimate from Saiki et al [53] with the ones from Gow et al [55] for the *ACE* expression. The resulting pooled estimate for the fold-change was 0.115 with a 95% CI=(−0.067;0.297) (Figure 3B). The same pooling of estimates was done for *ACE2* expression but using the estimates from Gow et al [55] and the reported mean log_2_ fold change reported by Smith et al [56]. The pooled estimate was 0.074 with a 95% CI=(−0.015;0.163). Finally, the remaining array-based GES studies did not report any evidence for differentially expressed *ACE*/*ACE2* genes between patients and healthy controls.

### 3.3 Analysis of *ACE/ACE2* gene expression in PBMCs among German participants

To consolidate evidence from previously published studies, gene expression levels of *ACE* and *ACE2* were quantified in PBMCs from 37 female patients diagnosed with ME/CFS (mean age = 41.1 years old) and 34 female healthy individuals (mean age = 37.4 years old) (Table 3). Patients and healthy participants were age-matched (Kolmogorov-Smirnov test, p=0.38). Patients had an average disease duration of 5.4 months (range = 0–24) with four of them without information about this variable.

As expected from PBMC samples, there was a higher mRNA level of *ACE* than of *ACE2* (Table 4, Figure 4A). Further analysis of the transformed expression did not present any significant correlation between *ACE* and *ACE2* expression levels (Spearman’s correlation coefficient = −0.120) (Figure 4B). Finally, linear regression models adjusted for age did not find any significant difference between patients and healthy controls (Table 4).

**Table 4.** Analysis of the linear regression models for the Box-Cox-transformed *ACE* and *ACE2* mRNA levels where data were only available for 26 ME/CFS patients.

| Analysis | Estimate (SE) | P-value |
| --- | --- | --- |
| Box-Cox transformed ACE |  |  |
| (Intercept) | 0.541 (0.032) | <0.001 |
| Age | 0.001 (0.001) | 0.328 |
| Disease Status (ME/CFS) | -0.013 (0.018) | 0.481 |
| Box-Cox transformed ACE2 |  |  |
| (Intercept) | 0.307 (0.038) | <0.001 |
| Age | -0.001 (0.001) | 0.137 |
| Disease Status (ME/CFS) | -0.006 (0.021) | 0.789 |

**Figure 4.**
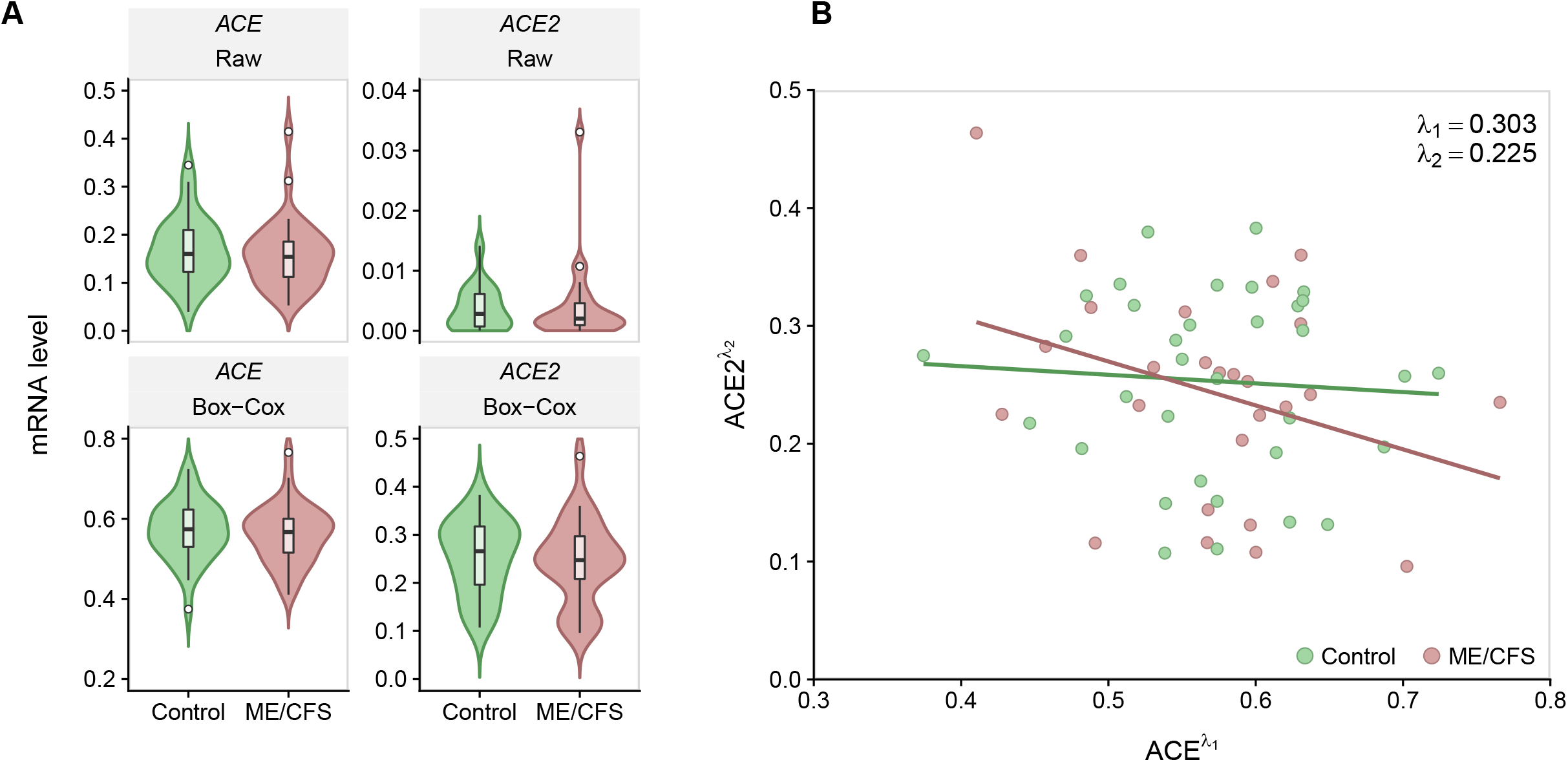
Analysis of *ACE* and *ACE2* gene expression from the German study. (**A)** Violin plots of *ACE* (left side) and *ACE2* (right side) mRNA raw data (upper row) and transformed data using the best Box-Cox transformation (lower row). Green-filled plots represent the cohort of healthy controls and rose-filled plots represent the ME/CFS-diagnosed patients. The best values for the Box-Cox transformation parameter λ are 0.303 and 0.225 for *ACE* and *ACE2* mRNA data, respectively. (**B)** Scatterplot between *ACE* and *ACE2* gene expression using the Box-Cox-transformed data (Spearman’s correlation coefficient = −0.120).

## 4. DISCUSSION

This research aimed to identify putative differences between patients with ME/CFS and healthy controls in terms of DNA methylation and gene expression of *ACE* and *ACE2*. Initially, we intended to perform similar research on current genome-wide association studies (GWAS) of ME/CFS [35,42,56,67,68]. However, we could not follow our original idea, because these studies did not make their data publicly available. With respect to these studies, evidence about the role of genetic factors on *ACE*/*ACE2* is inconclusive. Two studies reported many potential candidate SNPs for association with ME/CFS, but none of which was located in *ACE* and *ACE2* [56,67]. Other studies did not find any genetic marker across the genome to be associated with ME/CFS [35,42]. There was an additional GWAS reporting thousands of genetic associations with the disease [68], but this study did not perform all recommended quality controls [69].

With respect to published EWAS and GES, we could identify a female-specific association between ME/CFS and two potentially hypomethylated probes located in the TSS of *ACE* and *ACE2*, which suggests an increased expression of these genes in affected patients. However, our additional data set suggested that the gene expressions of *ACE* or *ACE2* in PBMCs were similar between patients and healthy controls. A similar conclusion was obtained from pooling estimates from different GES. These inconclusive results suggested that there is currently limited data to draw more conclusive inferences about the role of *ACE* and *ACE2* on ME/CFS. This data limitation is embodied not only in the small number of published studies, but also in the respective sample size used within each study, which limited the statistical power of the subsequent data analysis.

Current data limitation in ME/CFS can be explained by five main reasons. Firstly, there were only few EWAS and GES available in the literature. This limited number of studies could be related to a poor societal recognition of ME/CFS as a disease, which ultimately limits the funding available for the respective research. Access to limited research funding could also imply an additional difficulty in assembling multidisciplinary teams required to tackle the various challenging technical aspects of these studies.

Secondly, three published case-control GES based on microarray technology were excluded from this investigation, because they used broad or alternative case definitions of ME/CFS. Given the absence of an objective disease biomarker, the research community should aim to use consensual case definitions for research with the intention to make disease diagnostics comparable across studies while reducing heterogeneity between studies. In this regard, our requirement for ME/CFS diagnosis was the 1994 CDC/Fukuda definition or the 2003 CCC according to the recommendation for research given by the European Network on ME/CFS [18].

Thirdly, four RNA-seq studies were not included in our investigation due to unclear data quality. Issues concerning data quality is not an exclusive problem of ME/CFS studies, as highlighted by a comprehensive survey of the analytical steps taken by current RNA-seq studies [70]. In theory, there are several recommended steps for data processing and analysis for these studies [65]. In practice, different studies adopt distinct pipelines for data analysis with a possible impact on scientific reproducibility [70]. Again, a way to reduce between-study heterogeneity and to improve the respective data quality is to foster a stronger collaboration between ME/CFS researchers and bioinformaticians who have the technical competencies to conduct the correct processing of the data and the subsequent statistical analysis, as advised for the analysis of GWAS [69]. Notwithstanding the exclusion of the RNA-seq studies from this research, it is worth noting that none of them reported any difference between patients and healthy controls in terms of *ACE*/*ACE2* expression levels [61–64]. These findings are in agreement with the results obtained from our gene expression analysis for *ACE*/*ACE2* in German ME/CFS patients and healthy controls.

Fourthly, only a few of the published studies made their data publicly available. This issue was particularly limiting for a specific analysis of current GWAS, because none of these studies deposited the data in any open-access data repository. Currently, many funders and other science-related stakeholders are supporting the reuse and the long-term maintenance of scientific data generated by publicly funded research [71]. Above all, the benefit of a wide data-sharing practice is expected to accelerate scientific knowledge and to boost confidence in findings by allowing other researchers to take an alternative look at the same data. It could also promote collaboration among researchers, and to make science open to everyone, specifically, when it is funded by taxpayers and charities. Data sharing is also essential to cut down the costs of research by sharing resources among the research community. Reducing the costs of research by sharing limited resources is particularly important for the underfunded ME/CFS research field, as alluded above.

Fifthly, the re-analysis of publicly available data was based on small cohorts of patients and healthy controls. In general, a small sample size limits the statistical power to detect any hypothetical differences between patients with ME/CFS and healthy controls. This issue is particularly problematic for GWAS, EWAS and GES, whose statistical analysis typically involves the execution of thousands of association tests. In the case of GWAS, the number of association tests could even reach several million, as illustrated by Herrera et al [42]. Therefore, if correcting multiple testing is taken into account in the analysis, the most likely finding is the identification of relatively few disease associations, as demonstrated by Smith et al [56]. In the worst-case scenario, correcting for multiple testing in studies with small sample sizes leads to the absence of evidence for any disease association, as reported by different studies [35,42,72]. In the most optimistic scenario, we can hypothesize that *ACE* and *ACE2* are both genes whose genetic variation and gene expression profiles are at best moderately associated with ME/CFS. However, this prediction is yet to be confirmed with future studies investigating the specific role of these genes on patients with ME/CFS infected with SARS-CoV-2.

Given the high frequency of cardiovascular dysfunctions in patients with ME/CFS [24–29], it is also possible that the available DNA methylation and gene expression data could be biased towards study participants who were taking any medication to restore their normal cardiovascular function. In this regard, only one study excluded putative study participants taking any beta-blockers or ACE inhibitors [41]. Other studies excluded any potential participants with previous consumption of medications with immunomodulatory effects or with putative effects on epigenetic mechanisms [39,40,42], excluded any putative participant taking any regular medication [55], or reported that the healthy controls were free from any medication at the time of data collection [53]. One study conducted a review of current medications taken by the study participants [56]. However, it was unclear which medications were considered as exclusionary criteria.

Another cautionary note is that, for experimental convenience, gene expression and DNA methylation data sets were mostly derived from PBMCs and, as such, they may not reflect what occurs in nasal and pulmonary epithelial and endothelial cells, which are the main cellular targets of SARS-CoV-2 [73]. Interestingly, earlier studies on SARS-CoV-1 found the virus within T lymphocytes, macrophages, and monocyte-derived dendritic cells [74]. In the same line of evidence is the observation of lymphopenia in the blood of patients infected by SARS-CoV-2 [75,76]. It is then possible that SARS-CoV-2 also infects different immune cell subsets present in the blood. If so, the infection of PBMCs by this virus could open the door for a widespread of the infection to different organs. However, it is worth noting that, even if PBMCs are in fact infected by the SARS-CoV-2, it is unclear whether the virus uses the same invasion route via interaction with ACE2.

Previously, some authors hypothesized that patients with chronic conditions, such as those with hypertension, diabetes mellitus, or chronic obstructive respiratory disease, could be more susceptible to COVID-19 due to a putative upregulation of the *ACE2* gene [14]. A subsequent study could not confirm this hypothesis by analyzing the expression profiles of *ACE2* and other gene targets of SARS-CoV-2 in the lungs of these chronic patients [15]. However, this study failed to acknowledge a possible effect of the underlying genetic variation associated with *ACE2* in the respective results. In fact, a recent study showed a clear continental difference between different human populations based on *ACE2* polymorphisms alone [77]. Therefore, it is conceivable that different human populations could have a natural variation in the SARS-CoV-2 infectivity rate due to specific genetic variations in *ACE2* that can increase the binding affinity between ACE2 and the S1 protein encoded by SARS-CoV-2. In line with this view, a bioinformatic analysis suggested that specific *ACE2*-related SNPs are able to stabilize the interaction between ACE2 and the S1 protein of SARS-CoV-2 [78]. Given that genetic variation in *ACE2* is typically associated with cardiovascular diseases and there is currently no evidence for such genetic association with ME/CFS, we hypothesize that patients with ME/CFS have the same SARS-CoV-2 infectivity rate as any healthy individual on the basis of the *ACE2* data alone. On the other hand, it is known that patients with ME/CFS tend to have perturbations of the immune system with unresponsive natural killer cells upon antigen stimulation [79], defective B and T cell immune responses against the Epstein-Barr virus [80], decreased CD8+ T-cell cytotoxicity and activation [81], and increased percentage of regulatory T cells [82,83]. All of these clinical observations are possible reasons for frequent and persistent infections reported by some patients with ME/CFS [31]. Given all of these observations, a recent study suggested that the pathology of ME/CFS could be related to a hyper-regulated immune system via regulatory T cells [84]. As a corollary of this hypothesis, some patients with ME/CFS could have an increased SARS-CoV-2 infectivity rate not due to any underlying imbalanced expression of *ACE2*, but rather than due to a hypo-responsive (or hyper-regulated) immune system.

It is worth noting that the invasion of host cells by SARS-CoV-2 requires more than the simple interaction of the viral S1 protein with ACE2. Previously, it was found that SARS-CoV-1 interacts with the human transmembrane protease serine 2 (TMPRSS2) for its activation and its role of priming host cells for viral entry [85,86]. A similar interaction was hypothesized for SARS-CoV-2 infectivity [73]. In addition, TMPRSS2 is thought to induce SARS-CoV-1 cell entry through endocytosis via a mechanism of ACE2 cleavage, as reviewed elsewhere [87]. Similar mechanisms might occur in SARS-CoV-2 infections [4]. Another reported human protease potentially influencing SARS-CoV-2 infectivity is the A disintegrin and metallopeptidase domain 17 protein (ADAM17), which has an important role as a stress-response signal delivered to the immune system [88]. Like TMPRSS2, this protease coud also cleave ACE2, but with a different end-product [89]. As a consequence, the viral invasion seems less efficient in host cells whose ACE2 was preferentially cleaved by this protease than by TMPRSS2 [89]. At this moment, there is limited evidence for the role of these two proteases in the pathogenesis of ME/CFS. In this regard, one of the GES conducted a small gene expression study on different stress-response proteins including ADAM17 [53]. These authors did not find any significant difference in the expression of this protease between patients with ME/CFS and healthy controls. In addition, one of the EWAS provided evidence for hypomethylation of one ADAM17-related probe in patients with ME/CFS when compared to healthy controls [41]. Therefore, the analyses conducted here for ACE2 alone could serve as a guideline for future studies on these proteases related to SARS-CoV-2 infection. Dipeptidyl peptidase-4 (DPP4), also known as lymphocyte cell surface protein CD26, was found to be the main functional receptor for the host-cell entry by MERS [90,91]. This molecule is highly expressed in PBMCs including CD4+ and CD8+ T cells [8]. It is then possible that SARS-CoV-2 could infect PBMCs via a route involving DPP4 rather than ACE2. Interestingly, a study reported an increased proportion of natural killers and T cells expressing DPP-4/CD26+ in patients with CFS when compared to healthy controls [79]. A follow-up study confirmed this finding but also showed evidence for a decreased number of CD26 molecules in T lymphocytes and natural killer cells of patients with ME/CFS [92]. The same study suggested a decreased level of the soluble form of the molecule in the serum from patients. A similar observation was found in a recent study, but specifically for female patients whose disease was initiated after an infection [93]. Therefore, perturbations of the normal levels of DPP4 would appear to be a hallmark of ME/CFS pathogenesis. If DPP4 is indeed an alternative receptor for immune-cell invasion by SARS-CoV-2, specific research is needed to determine the infectivity rate of PBMCs from patients with ME/CFS. This would allow to determine the susceptibility of these patients to infections by SARS-CoV-2.

## 5. CONCLUSIONS

There is limited evidence for an altered expression of *ACE* and *ACE2* in PBMCs from patients affected by ME/CFS. At this stage, we could not rule out the hypothesis that patients and healthy controls alike could have the same infectivity rate of their PBMCs and other target cells by SARS-CoV-2. To investigate this hypothesis, further data should be analyzed, namely, on different human receptors (i.e., DPP4) that the virus can use to invade different host cells. In this regard, analyzing samples from the UK ME/CFS biobank [94,95] is a potential research avenue due to its large sample size, extensive clinical characterization of the respective study participants, and robust ethics.

## Data Availability

Data sets from epigenetic-wide and gene expression studies are publicly available at the NCBI GEO data repository. The data set from the German cohort is available from Prof Carmen Scheibenbogen upon request.

https://www.ncbi.nlm.nih.gov/geo/query/acc.cgi?acc=GSE59489

https://www.ncbi.nlm.nih.gov/geo/query/acc.cgi?acc=GSE93266

https://www.ncbi.nlm.nih.gov/geo/query/acc.cgi?acc=GSE156792

https://www.ncbi.nlm.nih.gov/geo/query/acc.cgi?acc=GSE111183

https://www.ncbi.nlm.nih.gov/geo/query/acc.cgi?acc=GSE14577

## Authors contributions

NS conceived this research. JM, AF, JCM, and NS performed the literature search. FW and JCM performed a comprehensive literature review about SARS-CoV-2/COVID-19 and their relationship with chronic conditions. FS, SB, HF, and CS were responsible for designing the study conducted in Charité, for recruiting the study participants, for collecting the blood samples, and processing them in the laboratory. JM, AF, CC, and NS performed the statistical analysis. JM, FS, AF, AG, LG, CS, EML, LN, JCM, FW, and NS interpreted and discussed the results. FW created the graphical abstract of the manuscript. NS and JM wrote the paper. All authors have read, revised, and approved the final version of the manuscript.

## Acknowledgements

JM and AF were fully funded by FCT – Fundação para a Ciência e Tecnologia, Portugal (ref.grant: SFRH/BD/149758/2019 and SFRH/BD/147629/2019, respectively). NS and CC were partially funded by FCT – Fundação para a Ciência e a Tecnologia, Portugal (ref. grant: UIDB/00006/2020). LN and EML acknowledge the funding from the National Institute of Allergy and Infectious Diseases (NIAID) of the National Institutes of Health (NIH -Award Number: R01AI103629), and from the ME Association (Award number: PF8947) for their studies on ME/CFS. The content of this paper is solely the responsibility of the authors and does not necessarily represent the official views of the NIH. The funding agencies did not have any role in the designing, data collection, data analysis, interpretation or writing-up the present manuscript.

## Abbreviations

ACE and ACE2: Human angiotensin-converting enzymes 1 and 2, respectively
ADAM17: A disintegrin and metallopeptidase domain 17 protein
CCC: Canadian Consensus Criteria
1994 CDC/Fukuda: 1994 Centers for Diseases Control and Prevention Criteria
COVID-19: Coronavirus disease 2019
DPP4: Dipeptidyl peptidase-4
EWAS: epigenome-wide association study or (studies)
GEO: Gene Expression Omnibus
GES: gene expression study (or studies)
GWAS: Genome-wide association study (or studies)
ME/CFS: Myalgic encephalomyelitis/Chronic Fatigue Syndrome
MERS: middle east respiratory syndrome
NCBI: National Centre for Biotechnology Information
PBMC: peripheral blood mononuclear cell
S1: viral spike glycoprotein
SARS-CoV-1: severe acute respiratory syndrome coronavirus-1
SARS-CoV-1/-2: severe acute respiratory syndrome coronavirus-1/-2
SNP: single nucleotide polymorphism
TMPRSS2: transmembrane protease serine 2
TSS: transcription start site

