## Supplementary Material for "The SARS-CoV-2 receptor angiotensin-converting enzyme 2 (ACE2) in Myalgic Encephalomyelitis/Chronic Fatigue Syndrome: analysis of high-throughput epigenetic and gene expression studies"

**Supplementary Table 1**

Nineteen and eight CpG probes located in ACE and ACE2 and shared between Infinium HumanMethylation450K and Infinium HumanMethylationEPIC arrays by Illumina. The annotation was obtained from the packages “IlluminaHumanMethylation450kanno.ilmn12.hg19” and “IlluminaHumanMethylationEPICanno.ilm10b2.hg19” available from Bioconductor.

| **Gene (chr)** | **Probes** | **Position** | **Annotation** |
| --- | --- | --- | --- |
| ACE (17) | cg09920557 | 61553938 | TSS1500 |
|  | cg25054907 | 61553954 | TSS1500 |
|  | cg02131967 | 61554106 | TSS1500 |
|  | cg02440279 | 61554400 | TSS200 |
|  | cg02040921 | 61554411 | TSS200 |
|  | cg19354750 | 61554413 | TSS200 |
|  | cg05952120 | 61554416 | TSS200 |
|  | cg24877195 | 61554604 | 1stExon |
|  | cg06751221 | 61554929 | Body |
|  | cg02261408 | 61559061 | Body |
|  | cg19802564 | 61561470 | Body;TSS1500 |
|  | cg19826045 | 61561602 | Body;TSS1500 |
|  | cg21796427 | 61562170 | TSS200;Body |
|  | cg04199256 | 61562197 | Body;5'UTR;1stExon |
|  | cg21094739 | 61572645 | Body;Body |
|  | cg01489398 | 61574335 | Body;Body |
|  | cg21881537 | 61574411 | Body;Body |
|  | cg21657705 | 61574500 | Body;Body |
|  | cg10468385 | 61574744 | 3'UTR;3'UTR |
| ACE2 (X) | cg23232263 | 15579482 | 3'UTR |
|  | cg05039749 | 15583512 | Body |
|  | cg05748796 | 15619337 | 5'UTR |
|  | cg16734967 | 15620103 | 5'UTR |
|  | cg08559914 | 15620240 | TSS200 |
|  | cg18877734 | 15621084 | TSS1500 |
|  | cg21598868 | 15621167 | TSS1500 |
|  | cg18458833 | 15621477 | TSS1500 |

**Supplementary Table 2**

Summary data of the CpG probes including SNP or coincided with a polymorphic SNP.

| **Annotation** | **Probe ID** | **Chromosome** | **Position** | **SNP ID** | **Ref allele/Alt Allele** | **Minor allele frequency** | |
| --- | --- | --- | --- | --- | --- | --- | --- |
|  |  |  |  |  |  | **North America** | **Europe** |
| SNP within probes | cg04199256 | 17 | 61562197 | rs191697444 | C/T | 0.00 | 0.00 |
|  | cg04199256 | 17 | 61562197 | rs12720723 | G/A | 0.12 | <0.01 |
|  | cg21881537 | 17 | 61574411 | rs117135474 | C/T | 0.00 | 0.00 |
|  | cg21881537 | 17 | 61574411 | rs200695691 | TTGCCC/T | 0.03 | 0.00 |
|  | cg10468385 | 17 | 61574744 | rs4365 | G/A | 0.00 | 0.04 |
|  | cg02481451 | 17 | 61594924 | rs149678437 | C/T | <0.01 | <0.01 |
|  | cg16734967 | X | 15620103 | rs182809041 | A/C | <0.01 | 0.00 |
| Polymorphic SNP | cg08559914 | X | 15620240 | rs186143966 | C/T | 0.00 | 0.00 |

**Supplementary Table 3**

Estimates of the best linear regression models for 5 significant CpG probes shown in Figure 1C. Healthy controls and the study of de Vega et al (2014) were considered the reference effects of the disease status and of the study indicator variable, respectively. The respective data are shown in Figure 2.

| **Analysis** | **Coefficient** | ***ACE*** | | | | | | | | ***ACE2*** | |
| --- | --- | --- | --- | --- | --- | --- | --- | --- | --- | --- | --- |
|  |  | **cg09920557** | | **cg19802564** | | **cg21094739** | | **cg10468385** | | **cg08559914^b^** | |
|  |  | **Estimate (SE)** | **P-value** | **Estimate (SE)** | **P-value** | **Estimate (SE)** | **P-value** | **Estimate (SE)** | **P-value** | **Estimate (SE)** | **P-value** |
| Overall | Intercept | -3.769 (0.065) | <0.001 | 1.551  (0.081) | <0.001 | 1.938 (0.096) | <0.001 | 3.082  (0.100) | <0.001 | 2.03547 (0.07766) | <0.001 |
|  | Disease status (ME/CFS) | -0.060 (0.092) | 0.519 | -0.016 (0.115) | 0.889 | -0.064 (0.136) | 0.638 | -0.118 (0.142) | 0.409 | -0.141 (0.048) | 0.004 |
|  | Study - de Vega et al (2016) | -2.367 (0.079) | <0.001 | 0.7071  (0.098) | <0.001 | 1.419 (0.116) | <0.001 | 2.437 (0.121) | <0.001 | 1.70315 (0.08513) | <0.001 |
|  | Study - Trivedi et al (2018)^a^ | 0.145 (0.092) | 0.117 | NA (NA) | NA | NA (NA) | NA | -0.625 (0.142) | <0.001 | -0.88672 (0.10334) | <0.001 |
|  | Study - Herrera et al (2018) | -2.245 (0.073) | <0.001 | 0.425 (0.091) | <0.001 | 0.740 (0.107) | <0.001 | 1.772 (0.112) | <0.001 | 1.27032 (0.08159) | <0.001 |
|  | Disease status (ME/CFS) × Study - de Vega et al (2016) | 0.131 (0.108) | 0.224 | -0.107 (0.133) | 0.425 | -0.198 (0.158) | 0.210 | -0.159 (0.165) | 0.336 | NA (NA) | NA |
|  | Disease status (ME/CFS) × Study - Trivedi et al (2018) | -0.315 (0.130) | 0.016 | NA (NA) | NA | NA (NA) | NA | 0.264 (0.199) | 0.186 | NA (NA) | NA |
|  | Disease status (ME/CFS) × Study - Herrera et al (2018) | 0.048 (0.102) | 0.638 | 0.072 (0.127) | 0.571 | 0.157 (0.150) | 0.295 | 0.181 (0.157) | 0.250 | NA (NA) | NA |
| Females | Intercept | -3.769 (0.068) | <0.001 | 1.406 (0.107) | <0.001 | 1.938 (0.093) | <0.001 | 3.082 (0.092) | <0.001 | 2.035 (0.068) | <0.001 |
|  | Disease status (ME/CFS) | -0.060 (0.096) | 0.535 | -0.224 (0.152) | 0.143 | -0.064 (0.132) | 0.629 | -0.117 (0.130) | 0.366 | -0.141 (0.045) | 0.002 |
|  | Study - de Vega et al (2016) | -2.367 (0.082) | <0.001 | 0.499 (0.130) | <0.001 | 1.419 (0.113) | <0.001 | 2.437 (0.111) | <0.001 | 1.703 (0.075) | <0.001 |
|  | Study - Trivedi et al (2018)^a^ | 0.145 (0.096) | 0.131 | NA (NA) | NA | NA (NA) | NA | -0.625 (0.130) | <0.001 | -0.887 (0.090) | <0.001 |
|  | Study - Herrera et al (2018) | -2.238 (0.079) | <0.001 | -0.233 (0.124) | 0.062 | 0.770 (0.108) | <0.001 | 1.786 (0.106) | <0.001 | 1.379 (0.073) | <0.001 |
|  | Disease status (ME/CFS) × Study - de Vega et al (2016) | 0.131 (0.112) | 0.241 | -0.112 (0.177) | 0.526 | -0.198 (0.153) | 0.198 | -0.159 (0.151) | 0.293 | NA (NA) | NA |
|  | Disease status (ME/CFS) × Study - Trivedi et al (2018) | -0.315 (0.134) | 0.020 | NA (NA) | NA | NA (NA) | NA | 0.264 (0.182) | 0.148 | NA (NA) | NA |
|  | Disease status (ME/CFS) × Study - Herrera et al (2018) | 0.029 (0.109) | 0.791 | 0.258 (0.173) | 0.138 | 0.166 (0.150) | 0.270 | 0.241 (0.148) | 0.104 | NA (NA) | NA |

^a^ The study of Trivedi et al (2018) discarded cg19802564 and cg21094739 from their analysis and, therefore, the respective data were not available in the NCBI GEO data repository.

^b^ The best regression model for cg08559914 included the main effects of disease status and study indicator covariate.
